# Interferon-activated neutrophils characterize coronary microvascular dysfunction in women with ANOCA

**DOI:** 10.64898/2026.10.30.26366961

**Authors:** E.A.M. de Jong, Daniek M.C. Kapteijn, Mark A. Daniels, Berber Zweers, Denise A.M. Peeters, Nienke Vrisekoop, Mete Civelek, Saskia Haitjema, Michal Mokry, Peter Damman, Tim P. van de Hoef, Ernest Diez Benavente, Hester M. den Ruijter

**Affiliations:** Laboratory of Experimental Cardiology, University Medical Center Utrecht, Utrecht University, Utrecht, the Netherlands; Respiratory Medicine, University Medical Center Utrecht, Utrecht University, Utrecht, the Netherlands; Center of Translational Immunology, University Medical Center Utrecht, Utrecht University, Utrecht, the Netherlands; Department of Cardiology, RadboudUMC, Radboud University, Nijmegen, the Netherlands; Department of Anesthesiology and Perioperative Medicine, University of California, Los Angeles, CA, USA; Central Diagnostic Laboratory, University Medical Center Utrecht, Utrecht University, Utrecht, The Netherlands; Department of Cardiology, Division Heart and Lungs, University Medical Center Utrecht, Utrecht University, Utrecht, the Netherlands

## Abstract

**Background:** Coronary microvascular dysfunction (CMD) is a common cause of ischemic symptoms in patients with angina and non-obstructive coronary arteries (ANOCA), predominantly women. Neutrophils may contribute to CMD via vascular inflammation and endothelial injury. We therefore investigated neutrophil and coronary endothelial cells in women with ANOCA.

**Methods:** In this cross-sectional study, we collected blood samples from ANOCA patients (n=123, 84% women) undergoing coronary function testing. Complete blood counts (CELL-DYN, n=123) and neutrophil activation (AQUIOS, n=76) were assessed. Isolated neutrophils were profiled using single-cell RNA-seq (n=11). Guidewire-derived coronary endothelial cells (n=7) isolated during coronary function testing were cultured to passage 2 for bulk RNA-seq.

**Results:** Of 103 women with ANOCA, 23 (22%) were diagnosed with CMD (CFR<2.5). These women showed higher NT-proBNP than women without CMD (234 [136-416] pg/ml vs. 112 [79-200] pg/ml, p<0.001). Neutrophil counts, neutrophil-to-lymphocyte ratio and systemic immune-inflammation index were higher among women with a lower coronary flow reserve (CFR). The proportion of activated neutrophils (CD62L^dim^) was inversely associated with CFR at baseline. Single-cell RNA-seq identified four neutrophil clusters, including interferon (IFN)-activated and immature neutrophils. The IFN-activated cluster was expanded in patients with lower CFR and its proportion was inversely associated with surface CD62L expression. Immature neutrophils showed reduced translational and increased inflammatory gene expression in patients with low CFR. Coronary endothelial cells from patients with increased CD62L^dim^ neutrophils showed higher proliferative/metabolic and lower IFN, complement and coagulation pathway activity.

**Conclusion:** Reduced CFR in women with ANOCA is associated with neutrophil activation, expansion of an IFN-activated neutrophil population, and altered immature neutrophil transcriptional programs. Neutrophil activation is accompanied by distinct endothelial signatures. These findings implicate neutrophil–endothelial interactions in CMD and identify IFN-associated neutrophil states as targets for mechanistic investigation.

**Clinical Perspective:** *What is new?:*

- This study provides the first characterization of circulating neutrophils in coronary microvascular dysfunction (CMD), identifying an increased proportion of interferon (IFN)-activated neutrophils and an IFN-enriched immature neutrophil phenotype
- Reduced neutrophil surface CD62L expression was associated with altered endothelial gene expression and increased endothelial proliferative and metabolic activity, suggesting a potential neutrophil-endothelial interaction in CMD

*What are the clinical implications?:*

- Neutrophil activation and IFN signaling emerge as candidate pathways for further investigation into the mechanisms underlying CMD and as potential targets for future therapeutic research
- The association between lower CFR in women and an increased IFN-activated neutrophil population supports further investigation of sex-specific immune mechanisms in CMD.

## Introduction

Angina with non-obstructive coronary arteries (ANOCA) is a highly prevalent yet clinically challenging condition among patients undergoing coronary angiography, particularly among women^1,2^. A central mechanism underlying ANOCA is coronary microvascular dysfunction (CMD), which is characterized by impaired regulation of coronary microvascular function, resulting in an inadequate increase in myocardial blood supply in response to myocardial demand^3^. Importantly, CMD is associated with high angina burden and adverse cardiovascular outcomes, including an increased risk of heart failure with preserved ejection fraction^4,5^. Despite this burden, effective and mechanism-guided treatments remain limited^6,7^. Elucidating the underlying pathophysiological mechanisms is therefore crucial to identify potential therapeutic targets and improve the management of patients with CMD.

Although the mechanisms underlying CMD remain incompletely understood, both coronary endothelial dysfunction and systemic low-grade inflammation have been implicated in its pathophysiology^8,9^. For instance, circulating IL-6 and TNF-α have been associated with reduced coronary flow reserve (CFR) in obese patients without clinical heart disease^10^. In addition, microvascular diseases are highly prevalent in chronic inflammatory diseases, including rheumatoid arthritis and systemic lupus erythematosus^11–14^, both of which disproportionately affect women.

Within these diseases, neutrophils play a central role^15–17^. They are key effector cells of the innate immune system and may link systemic inflammation to coronary microvascular impairment, as hypothesized previously in ANOCA^18^. Upon activation, neutrophils adhere to the endothelium and release reactive oxygen species, proteases, and inflammatory mediators, which can disrupt the endothelial barrier and impair nitric oxide mediated vasodilation. Neutrophil extracellular traps (NETs) may further contribute to endothelial injury by promoting endothelial activation, inducing oxidative stress and creating a prothrombotic environment^19,20^. Beyond their established role in inflammatory and autoimmune diseases, neutrophil activation has also been implicated in coronary plaque erosion^21^, a pathology that is also characterized by sex-differences^22^. These mechanisms provide a biological rationale for investigating the role of neutrophils in CMD.

Given the involvement of neutrophils in several female-predominant diseases, we investigated neutrophil abundance, activation and transcriptional states, together with coronary endothelial cell transcriptional profiles, in relation to CMD and invasively measured CFR in women with ANOCA.

## Methods

### 1. Study design and population

Blood sampling and isolation of coronary-catheterization derived endothelial cells was performed at the University Medical Center Utrecht as part of the ENDOTYPE biobank (TCbio 24-203). Written informed consent was obtained from all participants prior to the coronary function test. Participation in the Netherlands Registry of Invasive Coronary Vasomotor Function Testing (NL-CFT, 21-473)^23^ was a prerequisite for inclusion in the ENDOTYPE biobank, which enabled linkage of the biological samples to the corresponding clinical and procedural data. The ENDOTYPE biobank was approved by the Biobank Committee operating under the responsibility of the Medical Ethical Committee of the University Medical Center Utrecht. All procedures were conducted in accordance with the Declaration of Helsinki. Data supporting the findings of this study are available from the corresponding author upon reasonable request.

For the present study, patients with obstructive coronary artery disease, as identified prior to the coronary function test and patients with incomplete CFR measurements were excluded.

### 2. Diagnostic coronary function testing

After exclusion of functionally significant epicardial coronary artery disease (non-hyperemic pressure ratio >0.89 or fractional flow reserve >0.80), ANOCA patients underwent invasive coronary function testing in the left anterior descending artery, including vasospasm provocation testing and coronary vasodilatory capacity assessment. The procedure has been described in detail previously^24^. Intracoronary acetylcholine provocation testing was used to assess coronary vasospasm, using incremental acetylcholine doses of 20, 100 and 200mg. Spasm diagnosis was based on the Coronary Vasomotor Disorders International Study Group criteria^25^: epicardial spasm required reproducible angina, ischemic electrocardiographic changes, and >90% epicardial vasoconstriction; microvascular spasm required recognizable anginal symptoms and ischemic electrocardiographic changes without >90% epicardial vasoconstriction. Tests with symptoms or ischemic electrocardiographic changes alone were considered inconclusive (or equivocal).

Coronary vasodilatory capacity was assessed using continuous thermodilution, using a pressure- and temperature-sensitive sensor equipped guidewire (PressureWire X, Abbott Vascular, Santa Clara, USA) together with a RayFlow infusion catheter (Hexacath, Paris, France) positioned in the proximal left anterior descending artery, as described previously^24^. Absolute coronary blood flow was quantified under resting and hyperemic conditions during saline infusion at 10 and 20 mL/min, respectively. Measurements were obtained using dedicated software (CoroVentis, Uppsala, Sweden) based on the continuous thermodilution principle. Absolute microvascular resistance was calculated as distal coronary pressure divided by absolute coronary blood flow. Coronary flow reserve (CFR) was defined as the ratio of hyperemic to resting absolute coronary blood flow and microvascular resistance reserve was calculated as previously described^26^. For the present study, a diagnosis of CMD was defined as an absolute CFR <2.5, consistent with established diagnostic thresholds for impaired vasodilatory capacity^27^.

### 3. Blood withdrawal

Blood samples were collected from ANOCA patients during the coronary function test, before any intracoronary measurements were performed. First, systemic blood samples were obtained from the radial or femoral arterial sheath immediately after vascular access was established. Second, local blood samples were collected from the guiding catheter after engagement of the left anterior descending artery. Both blood samples were collected following heparin administration to minimize procedural influences.

### 4. Clinical biomarker measurements

Clinical biomarkers, including total cholesterol, low-density lipoprotein cholesterol, high-density lipoprotein cholesterol, triglycerides, creatinine and N-terminal pro-B-type natriuretic peptide (NT-proBNP), were measured retrospectively in plasma samples obtained from systemically collected EDTA-anticoagulated blood. Blood samples were centrifuged at 1,850*g* for 10 minutes at room temperature with brake (Hettich centrifuge). Afterwards, the plasma was stored at −80°C and was thawed once before clinical biomarker measurements.

Total cholesterol, low-density lipoprotein cholesterol, high-density lipoprotein cholesterol, triglycerides and creatinine were measured using the Atellica CI analyzer (Siemens Healthineers, Forchheim, Germany) and NT-proBNP was measured using the Atellica IM immunoassay analyzer (Siemens Healthineers). Lipid parameters and creatinine were quantified using enzymatic assays with photometric detection. NT-proBNP was quantified using a fully automated, two-site sandwich immunoassay based on direct chemiluminescence.

### 5. Routine hematological complete blood cell counts

Hematological complete blood cell counts were routinely measured in systemic and local EDTA-anticoagulated blood samples using the CELL-DYN Sapphire hematology analyzer (Abbott Diagnostics, Abbott Park, IL, USA). Available measurements for all included patients were retrieved from the Utrecht Patient Orientated Database (UPOD)^28^. Parameters included red blood cell indices (hemoglobin, hematocrit, red blood cell count, mean corpuscular volume, mean corpuscular hemoglobin concentration and red cell distribution width), platelet count, total leukocyte count, and differential leukocyte counts (neutrophils, lymphocytes, monocytes, eosinophils and basophils). The neutrophil-to-lymphocyte ratio (NLR), monocyte-to-lymphocyte ratio (MLR), and systemic immune-inflammation index (SII, platelet x neutrophil/lymphocyte ratio) were calculated from absolute counts.

### 6. AQUIOS flow cytometry and data analysis

Immediately after collection, systemic and local blood samples collected in sodium-heparin tubes, were loaded onto the AQUIOS CL ‘Load & Go’ flow cytometer (Beckman Coulter, Miami, FL, USA)^29^. This platform fully automates sample preparation and flow cytometric analysis, including sample mixing, pipetting and staining with antibodies. After a 15-minute incubation, red blood cells were lysed by adding 335 ml of lysing reagent A. Next, 100 ml of lysing reagent B was added to stop the lysis and preserve leukocyte integrity. The prepared samples were then aspirated for analysis. Absolute leukocyte counts were determined by electronic volume measurement.

To assess neutrophil surface marker expression, a custom antibody panel was used, consisting of CD64-PC7 (clone 22, Beckman Coulter, Miami, FL, USA), CD16-FITC (clone 3G8, Beckman Coulter), CD11b-PE (clone Bear1, Beckman Coulter), CD62L-ECD (clone DREG56, Beckman Coulter) and CD10-PC5 (clone ALB1, Beckman Coulter). To assess neutrophil responsiveness, each sample was measured both unstimulated and after stimulation with the bacterial/mitochondrial-derived stimulus N-formyl-norleucyl-leucyl-phenalalanine (fNLF, BioCat GmbH, Heidelberg, Germany) at a final concentration of 10^-5^ M. fNLF is a bacterial formyl-peptide receptor agonist that stimulates neutrophil activation, including degranulation and the mobilization of adhesion molecules^30^.

For detailed analysis, data files were exported from the AQUIOS CL flow cytometer and analyzed using FlowJo software (Tree Star Inc. Ashland, OR, USA). Granulocytes were gated based on forward and side scatter, followed by neutrophil identification based on CD16 and CD11b co-expression. The number of cells/L was calculated by multiplying the gated events with a correction factor of 232558.139535. The median fluorescent intensity (MFI) of the different markers was determined within this neutrophil gate. Neutrophil phenotypes were classified by quadrant gating based on CD16 and CD62L expression, as previously described^31^. The gating strategy for CD16/CD62L-based phenotyping is shown in **Supplemental Figure S1**.

### 7. Neutrophil isolation and single-cell library preparation

Immediately after blood collection, polymorphonuclear neutrophils were isolated by layering of EDTA-anticoagulated systemic whole blood onto of Polymorph prep™ (Axis-Shield, CAT#1114683; 1:1 v/v), followed by immediate centrifugation at 500g for 35 min at room temperature, as described in detail before^21^. Leftover red blood cells were removed via hypotonic lysis using 10 mL of ice-cold dH₂O for 10-30 seconds, immediately neutralized with 10 mL of 2xPBS (Sigma Aldrich, CAT#D8537), and centrifuged at 400g for 5 minutes at 4°C. For single-cell RNA sequencing, a 100 μL aliquot of the purified PMN suspension (diluted with 1x PBS to 3.5 × 10⁵ cells/mL) was processed according to the GEXSCOPE Single-Cell RNA Library for NEO Handbook^32^.

### 8. Library QC and sequencing of single-cell RNA libraries of isolated neutrophils

The library concentration and fragment size distribution were assessed using a Qubit High Sensitivity dsDNA kit on a Qubit 4 fluorometer (ThermoFisher Scientific, Waltham, MA, USA) and the TapeStation High Sensitivity D1000 tape (Agilent Technologies, Santa Clara, CA, USA). The libraries were pooled in an equimolar fashion and sent to GeneWiz (Azenta Life Sciences, Leipzig, Germany) to be sequenced on an Illumina NovaSeq X to an average depth of 30,000 reads/cell in a 2×150 bp manner. The BCL files were demultiplexed to fastq files on Illumina’s BaseSpace platform. Libraries were excluded from downstream analysis when sequencing quality control indicated a low median UMI count per cell, consistent with low sample or cDNA quality.

### 9. Single-cell data analysis

Raw sequencing fastq files were processed into gene expression matrices using CeleScope™ version 2.11.3 (Singleron Biotechnologies, Cologne, Germany, https://github.com/singleron-RD/CeleScope). Fastq file barcode chemistry determination and barcode correction were performed before being demultiplexed and gene counts generated using the STARSolo single-cell pipeline with STAR version 2.7.11a. The human reference genome (GRCh38) was used for mapping the reads and annotation was done with Ensembl version 99. Cell calling was based on bimodal modeling using the EmptyDrops algorithm. Samples were additionally excluded from downstream analysis if their gene-barcode matrices were dominated by near-empty droplets, with the majority of nominally called cells failing minimum UMI/gene-count thresholds upon reanalysis.

Doublet detection and standard per-cell quality-control filtering (based on library size, gene number and mitochondrial and hemoglobin content) were performed per sample prior to merging the individual patient libraries into a single dataset for downstream integration. Non-neutrophil populations were excluded based on marker gene expression. Data were normalized using SCTransform (glmGamPoi), followed by principal component analysis (30 PCs), batch correction using Harmony, and shared nearest-neighbor clustering. A clustering resolution of 0.1, yielding four clusters, was selected based on mean silhouette width. Clusters were annotated using canonical markers, differential gene expression analysis (Wilcoxon rank-sum test; log_2_ fold change threshold 0.25) and pathway enrichment (clusterProfiler; GO Biological Process). To assess concordance with an independent human neutrophil reference atlas, per-cluster module scores were calculated (AddModuleScore, Seurat) using neutrophil-specific gene signatures derived from a large single-cell atlas of human neutrophils^33^.

Transcriptional relationships among clusters were further characterized using trajectory and pseudotime analysis. Trajectory inference was performed using Slingshot^34^. The trajectory was rooted in the immature/banded cluster, based on its high ribosomal gene content, indicating a less mature neutrophil state. Pseudotime values were extracted separately for each lineage. Cells not assigned to a given lineage received no pseudotime value for that lineage and were excluded from the corresponding analysis. To visualize gene expression dynamics along each trajectory, the top four marker genes per cluster were identified. For each gene and each lineage, expression (SCT-normalized) was fitted against pseudotime using local polynomial regression (loess, span=0.3) across 200 evenly spaced points spanning that lineage’s pseudotime range. The smoothed values were z-scored per gene within each lineage. To test whether this lineage bias was already detectable within the immature cluster itself, composite module scores representing the interferon-stimulated and aged transcriptional programs were computed. These were derived from genes differentially expressed between these two clusters (FindMarkers, Wilcoxon rank-sum test, log2 fold-change threshold 0.25, adjusted p<0.05), with genes enriched in each cluster scored separately (AddModuleScore). The difference between the two scores (IFN-vs-Aged score) was calculated per cell. Within the immature neutrophil cluster, this score was related to continuous CFR using linear mixed-effects model (lme4), with patient included as a random effect to account for multiple cells per patient and compared against an intercept-only null model by likelihood ratio test.

### 10. Endothelial cell isolation and bulk RNA sequencing

Endothelial cells were isolated from catheterization material obtained during the diagnostic coronary function test and cultured as described previously^24^. In brief, guidewires were cut into ±5 cm segments and transported on ice in complete endothelial cell medium. To release adherent endothelial cells, the segments were rotated for 10 minutes at 4°C, after which the segments were removed from the medium. Cells were pelleted by centrifugation, resuspended in complete endothelial cell medium and seeded onto a gelatin-coated 12-well plate. Cultures were maintained at 37°C and 5% CO2 with medium refreshed three times weekly. At passage 2, endothelial cells were harvested for bulk RNA sequencing, which was performed as described previously^24^.

### 11. Statistical analysis

Normally distributed continuous variables, as assessed by the Shapiro-Wilk test, are presented as mean ± SD, while non-normally distributed variables are shown as median [IQR]. Categorical variables are reported as number (%). Group comparisons (patients with vs. without a CMD diagnosis, defined as CFR<2.5) were performed using Welch’s t-test or the Mann-Whitney U test, as appropriate, and the chi-squared or Fisher’s exact test for categorical variables. Associations between continuous variables were assessed using Spearman’s rank correlation, with age-adjusted estimates obtained from linear regression models. Right-skewed variables (NT-proBNP, NLR, MLR, SII) were log-transformed prior to inclusion in linear regression models. When appropriate, continuous predictors were standardized to obtain effect estimates per SD with 95% confidence interval.

Differential gene expression analysis was performed using DESeq2 on patient-level aggregated (pseudobullk) counts for both the neutrophil single-cell RNA sequencing clusters and the coronary catheterization-derived endothelial cells. For neutrophil clusters, genes were retained for analysis if at least 3 patients had ≥5 counts and models were adjusted for mean nFeature and cell number per patient. For endothelial cells, DESeq2 modeled baseline CD62L MFI, measured by AQUIOS flow cytometry as continuous predictor (z-scored to obtain effect estimates per SD) adjusting for age. Genes were retained if at least 3 samples had ≥10 counts and log2 fold-change estimates were shrunk using the apeglm method. Across both analyses, genes with adjusted p<0.05 were considered significant, and volcano plots were generated using the EnhancedVolcano package. Gene set enrichment analysis was performed using fgsea on genes ranked by the DESeq2 Wald statistic, testing Hallmark gene sets. Pathways with Benjamini-Hochberg-adjusted p<0.05 were considered significant. For the present study, all tests were two-sided, with p<0.05 considered statistically significant unless otherwise specified. Analyses were performed in R (version 4.5.1).

## Results

### 1. Study population

In total, 128 ANOCA patients undergoing coronary function testing consented to participate in the ENDOTYPE biobank for withdrawal of both systemic and local coronary blood. Four patients (3%) were excluded due to obstructive coronary artery disease detected during the procedure, and in one patient (1%), coronary flow reserve could not be measured due to technical difficulties. Ultimately, blood samples were available from 123 ANOCA patients, of whom 84% were women. Given the focus on women in the present study, baseline characteristics of the women are presented in **Table 1**, while corresponding data of men are provided in **Supplemental Table S1.**

**Table 1.**
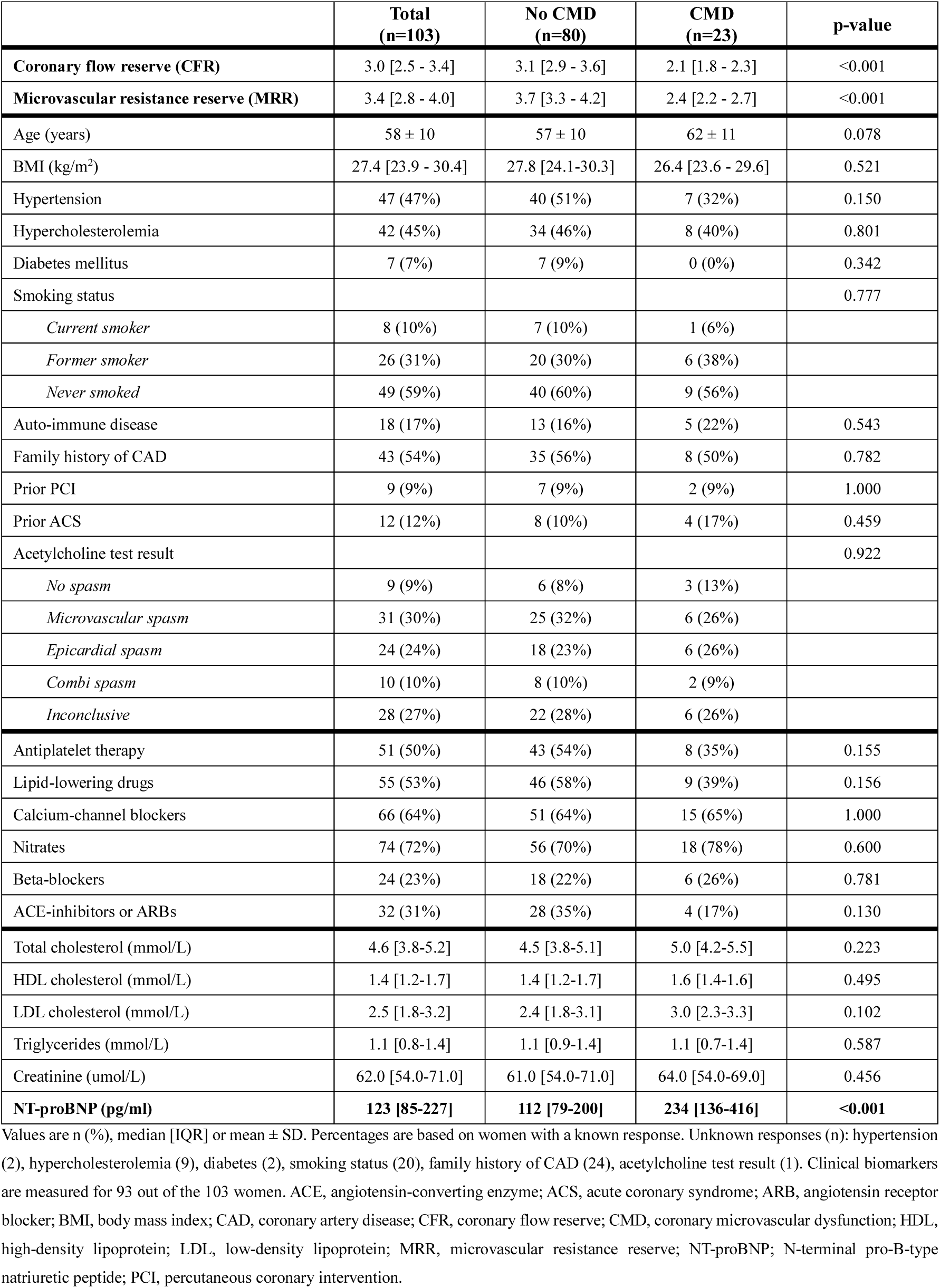
Baseline characteristics in women with ANOCA.

Among the women in the study, the mean age was 58 ± 10 years. Cardiovascular risk factors were common, including hypertension (47%) and hypercholesterolemia (45%), while 7% had diabetes mellitus and 17% were diagnosed with an auto-immune disease. Acetylcholine spasm provocation testing resulted in a diagnosis of epicardial spasm in 24 women (24%), microvascular spasm in 31 women (30%), and combined epicardial and microvascular spasm in 10 women (10%). An inconclusive spasm test (not meeting the COVADIS criteria) was observed in 28 women (27%), while 9 women (9%) had a negative spasm test. Twenty-three women (22%) had a CFR < 2.5 and were clinically diagnosed with CMD, while 80 women (78%) had a CFR ≥ 2.5. Median MRR was significantly lower in women with a CMD diagnosis (2.4 [2.2-2.7]) compared to women without a CMD diagnosis (3.7 [3.3-4.2]) (p<0.001). Baseline demographic characteristics, cardiovascular risk factors, biomarker profiles, comorbidities, and medication use did not differ significantly between women without CMD and with CMD (all p>0.10), with the exception of age, which showed a trend towards a higher age in women with CMD than those without CMD (62 ± 11 years vs. 57 ± 10 years, p=0.078) (**Table 1**).

Women with CMD are known to have a higher risk for developing heart failure^4^. Therefore, we measured plasma NT-proBNP concentrations. NT-proBNP concentrations were higher in women with CMD compared to those without CMD (234 [136-416] vs. 112 [79-200] pg/mL, p<0.001), and this remained significant after adjustment for age (β= −0.154, p<0.001). In addition, NT-proBNP inversely correlated with CFR (rho =-0.24, p=0.023) also when adjusted for age (β= −0.154, p=0.038).

### 2. Systemic neutrophil abundance and neutrophil-related inflammatory indices are inversely associated with CFR in women

First, to study the circulating neutrophil profile of women with ANOCA in this cohort, we assessed systemic complete blood counts (CELL-DYN, n=103) and flow-cytometry derived neutrophil counts (AQUIOS, n=67) in this population. CELL-DYN-derived absolute neutrophil counts were not different between women with and without a CMD diagnosis (4.30 [3.70 - 6.06] vs. 4.29 [3.44 - 5.17] x10^9^ cells/L, p =0.621). However, neutrophil count showed an inverse association with CFR after age adjustment (β= −0.217, p=0.022) (**Figure 1A**). AQUIOS-derived neutrophil counts showed a similar inverse association with CFR, although not statistically significant (rho =0.163, p=0.188; age-adjusted β =-0.204, p=0.082) (**Figure 1B**).

**Figure 1.**
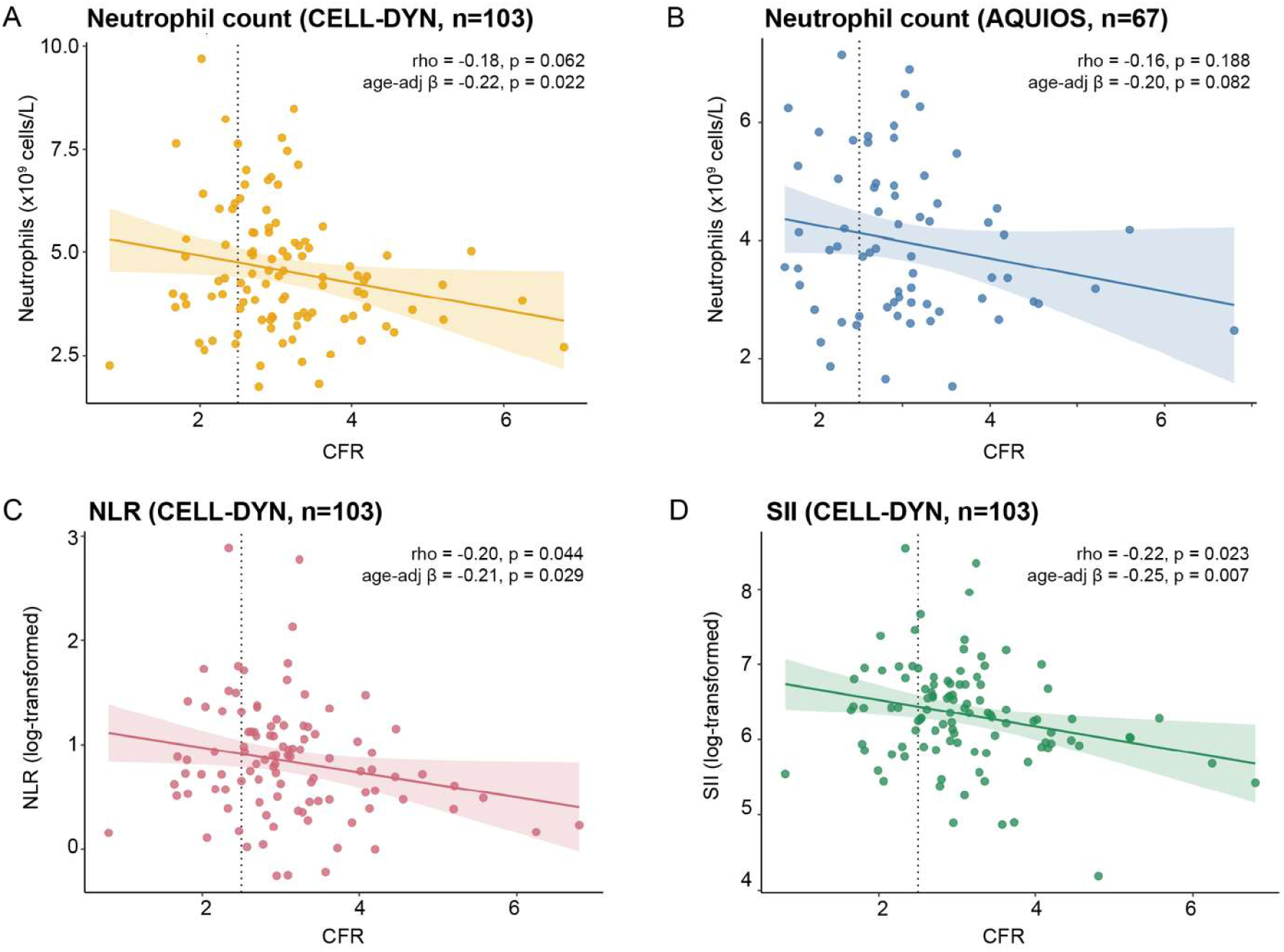
Neutrophil abundance and inflammatory indices in relation to coronary flow reserve (CFR) in women with angina and non-obstructive coronary arteries (ANOCA). Associations between systemic neutrophil parameters and CFR. (A) Absolute neutrophil count (CELL-DYN, n=103). (B) Neutrophil count measured by flow cytometry (AQUIOS; n=67). (C) Neutrophil-to-lymphocyte ratio (NLR, log-transformed; CELL-DYN, n=103). (D) Systemic immune-inflammation index (SII, log-transformed; CELL-DYN, n=103). Solid lines indicate linear regression fits with 95% confidence interval; the dotted vertical line marks the CFR<2.5 cutoff used for diagnosis of coronary microvascular dysfunction. Spearman’s rank correlation coefficient (rho) is reported for the unadjusted association with CFR; β denotes the standardized regression coefficient (per SD) from an age-adjusted (age-adj) linear model.

Clinically, systemic neutrophil-related inflammatory indices, such as the neutrophil-to-lymphocyte ratio (NLR) and systemic immune-inflammation index (SII, platelet x neutrophil/lymphocyte ratio) have been associated with coronary microvascular function, although findings have been inconsistent^35–38^. We therefore studied the NLR and the SII. The NLR (log-transformed) showed no difference between women with and without CMD (2.07 [1.74-4.02] vs. 2.26 [1.60-3.02], p=0.539) but was inversely associated with CFR (rho = −0.199, p=0.044; β= −0.206, p=0.029) (**Figure 1C**). Similarly, the SII (log-transformed) showed no difference between women with CMD and without CMD (614.6 [356.5 - 1025.6] vs. 532.9 [384.9 - 786.2], p=0.408), but a significant inverse correlation with CFR (rho =-0.223, p=0.023; age-adjusted β =-0.252, p=0.007) (**Figure 1D**).

In local blood samples, collected sequentially during the same catheterization procedure, women (n=94) with a lower CFR similarly had higher values for all three CELL-DYN indices and AQUIOS neutrophil counts, consistent with findings in systemic blood, although the effects were generally weaker (**Supplemental Table S2 and Supplemental Figure S2**). In contrast to the pattern observed in women, in the smaller cohort of men (CELL-DYN, n=20; AQUIOS, n=9), neutrophil-count parameters showed trends towards positive associations with CFR. However, these trends did not reach statistical significance in either systemic or local blood **(Supplemental Table S3 and Supplemental Figure S3)**

Beyond neutrophil-related measures, eosinophil counts in women showed positive associations with CFR in both systemic and local blood, although the systemic association was no longer significant after age adjustment. In contrast, the association in local blood remained significant after adjustment. Local hemoglobin levels also differed between women with and without CMD, but not after age adjustment (**Supplemental Table S2**).

Together, these findings suggest that in women with ANOCA, higher systemic neutrophil abundance and neutrophil-related inflammatory indices are associated with a lower CFR, independent of age. Next, we examined whether neutrophil characteristics beyond cell abundance, including cellular activation and phenotype, were associated with CFR.

### 3. Flow cytometry characterization of neutrophils links activated CD62L^dim^ neutrophils and CFR

To test the hypothesis that women with a low CFR have more activated neutrophils compared to women with a normal CFR, we tested neutrophil surface marker expression. We used flow cytometry (AQUIOS, n=67) to measure a panel of neutrophil activation markers, including CD62L, CD64, CD10, CD16, and CD11b. Both neutrophils isolated from systemic blood at baseline and after *ex vivo* stimulation with fNLF were characterized. CD62L is constitutively expressed on resting neutrophils and is rapidly shed from the cell surface upon activation^39^. We therefore used an established cutoff (as described in the Methods section) to quantify the proportion of CD62L^bright^ and CD62L^dim^ neutrophils, the latter reflecting a more activated neutrophil phenotype.

At baseline, a higher percentage of CD62L^dim^ neutrophils was associated with lower CFR (rho = −0.189, p=0.126; age-adjusted β=-0.274 per SD, p=0.018) (**Figure 2A and B**). Consistent with this, the MFI of CD62L showed the same trend towards a positive association with CFR (rho = 0.173, p=0.160; age-adjusted β = 0.196 per SD, p=0.094). The MFIs of CD64, CD10, CD16 and CD11b showed no association with CFR (all p>0.50) (**Figure 2A**).

**Figure 2.**
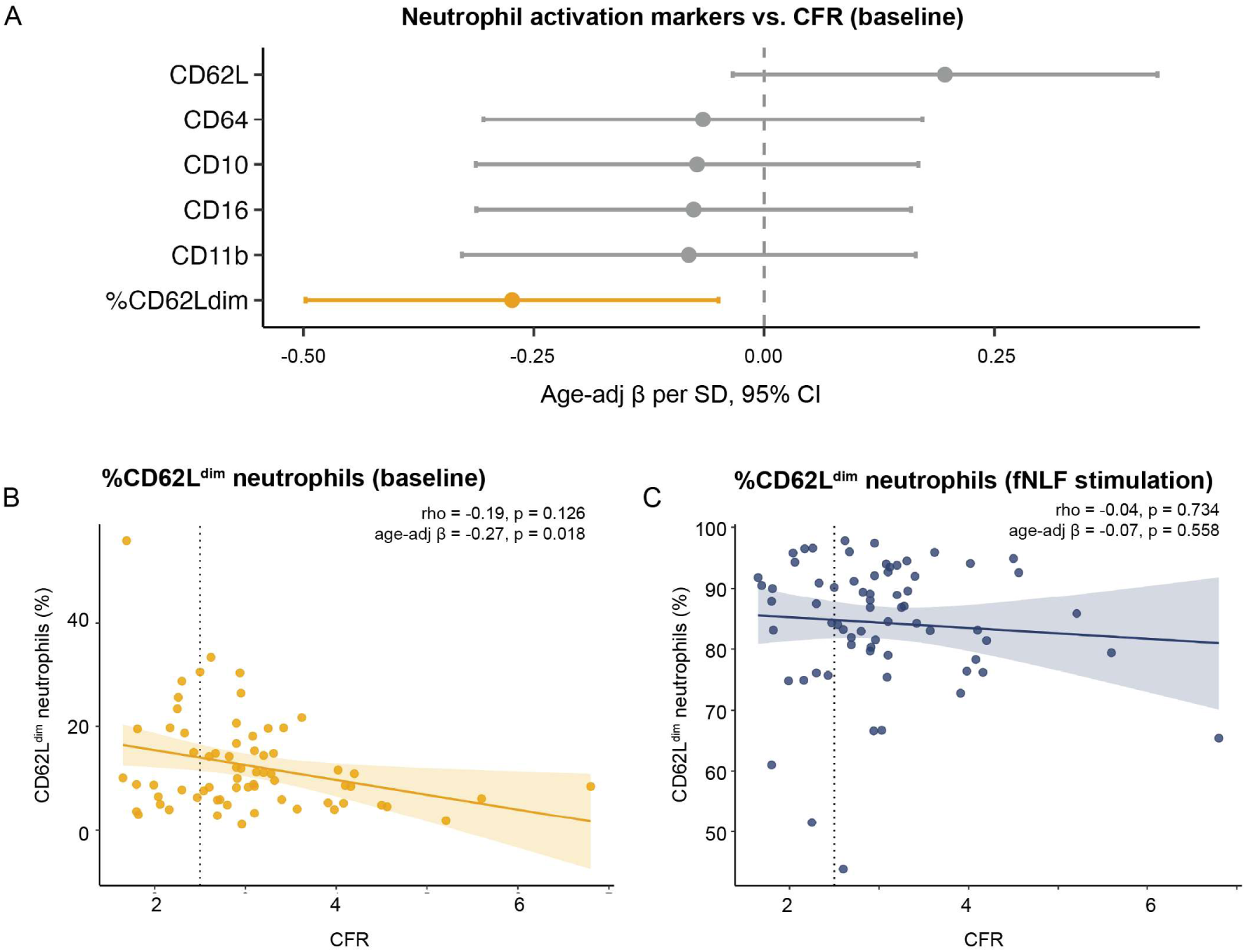
Neutrophil activation markers in relation to coronary flow reserve (CFR) in women with angina and non-obstructive coronary arteries (ANOCA). Systemic neutrophil activation marker expression (AQUIOS flow cytometry, n=67) in relation to CFR. (A) Forest plot of age-adjusted standardized regression coefficients (β per SD, 95% CI) for the association between CFR and median fluorescence intensity (MFI) of CD62L, CD64, CD10, CD16 and CD11b, and the percentage of CD62L^dim^ neutrophils (%CD62L^dim^), measured at baseline. Markers reaching statistical significance (p<0.05) are highlighted in orange. (B) %CD62L^dim^ neutrophils at baseline versus CFR. (C) %CD62L^dim^ neutrophils following ex vivo stimulation with fNLF versus CFR. In (B) and (C), the solid line shows the linear regression fit with 95% CI; the dotted vertical line marks the CFR<2.5 cutoff used to diagnose coronary microvascular dysfunction. Spearman’s rank correlation coefficient (rho) is reported for the unadjusted association with CFR; β denotes the standardized regression coefficient (per SD) from an age-adjusted (age-adj) linear model

To test whether this higher neutrophil activation in women with low CFR reflected a difference in baseline state or a difference in activation capacity, we assessed the percentage of CD62L^dim^ neutrophils after *ex vivo* stimulation with the bacterial peptide fNLF. The percentage of CD62L^dim^ neutrophils was no longer associated with CFR after stimulation (**Figure 2C**). This suggests that the association with CFR may reflect a baseline activation state of circulating neutrophils rather than differences in their response to stimulation.

In local coronary blood, the percentage of CD62L^dim^ neutrophils showed a similar, although weaker association with CFR **(Supplemental Figure S2**). In men (n=9), the association in local blood was in the opposite direction compared to women and reached statistical significance. In systemic blood, no significant association was observed, and the direction was inconsistent between unadjusted and age-adjusted models (**Supplemental Figure S3**).

Overall, lower CFR in women was associated with a higher percentage of CD62L^dim^ neutrophils at baseline, which suggests a link between reduced CFR and an altered neutrophil activation state.

### 4. Single-cell RNA sequencing reveals an interferon-activated neutrophil cluster associated with CMD and a reduced CFR

To characterize the transcriptional heterogeneity of circulating neutrophils, we performed single-cell RNA sequencing of isolated neutrophils from ANOCA patients (n=16, 12 women and 4 men). After quality control, neutrophils from 11 ANOCA patients (8 women and 3 men) contributed to a total of 55,915 cells that were analyzed. After exclusion of contaminating cell populations, the final dataset comprised 53,216 neutrophils from 11 patients (**Supplemental Table S4**), with a median of 4,747 cells per patient and 416 detected genes per cell.

Unsupervised clustering of the neutrophils identified four transcriptionally distinct circulating neutrophil clusters (**Figure 3A**). Based on marker gene expression and pathway enrichment, these clusters were annotated as 1) An immature cluster (*RPSA, RPS8, RPL22, RPS12*), 2) A mature cluster (*MMP9, RFLNB, PADI4, S100A12),* 3) An aged cluster (*SLPI, PI3, SIGLEC10, ADGRE2)* and 4) An interferon (IFN)-activated cluster (*RSAD2, ISG15, HERC5, IFIT1*) (**Figure 3B, Supplemental Table S5)**. Each neutrophil cluster was defined by a distinct transcriptional program (**Figure 3C, Supplemental Table S6**). The immature cluster was enriched for pathways involved in protein synthesis, consistent with a less mature neutrophil state. The mature cluster was enriched for leukocyte activation and chemotaxis, the aged cluster for ferroptosis and innate immune signaling, and the IFN-activated cluster for antiviral/interferon-response pathways. Comparison with an independent human neutrophil reference atlas^33^, showed concordant signatures for the mature, aged and IFN-activated cluster, but not for the immature cluster, which is instead characterized by high ribosomal gene expression rather than classic granule genes (**Supplemental Figure S4**).

**Figure 3.**
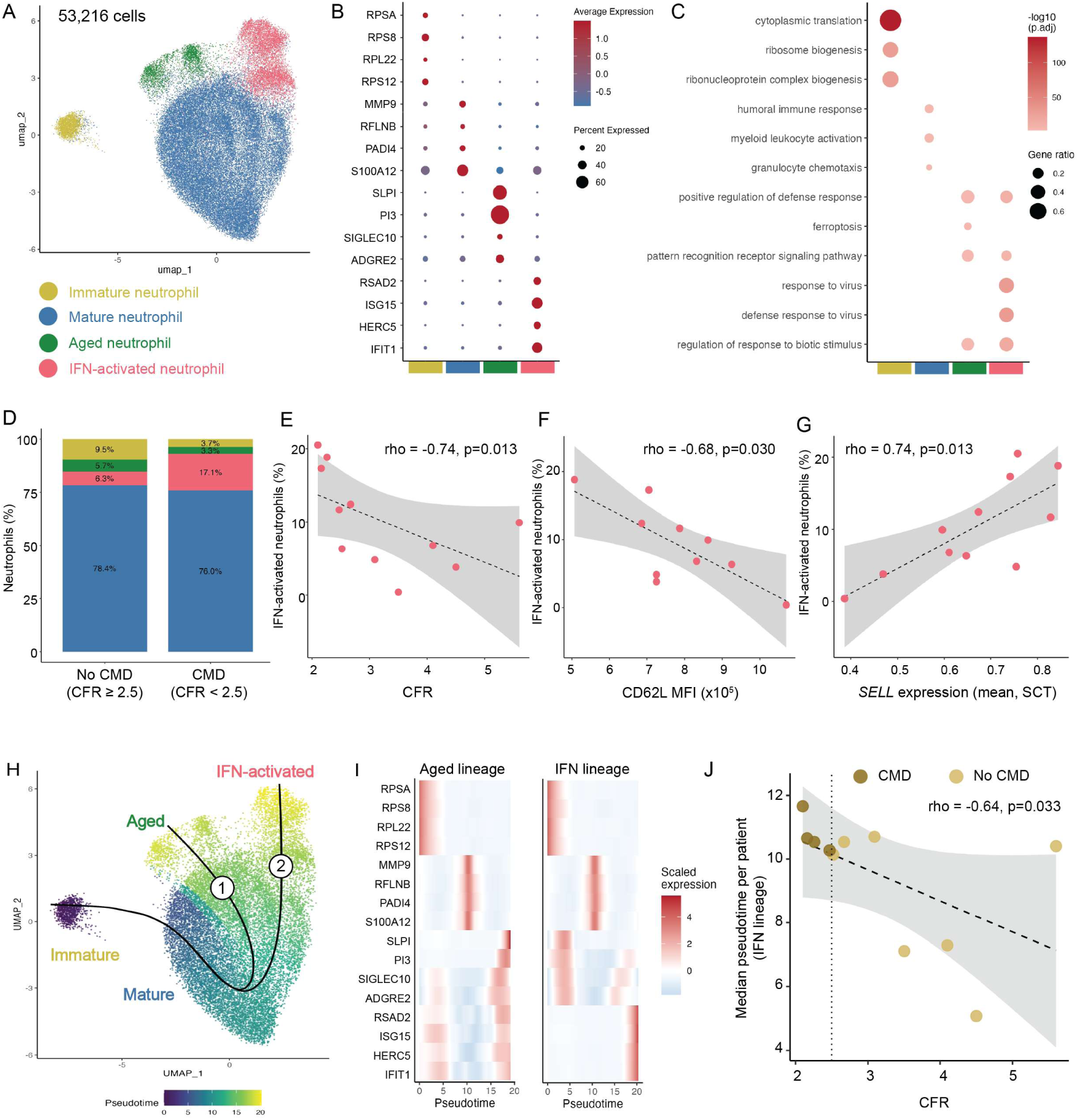
Single-cell transcriptional landscape and pseudotime trajectory of circulating neutrophils in patients with angina and non-obstructive coronary arteries (ANOCA). Single-cell RNA sequencing of circulating neutrophils (53,216 cells, 11 patients). (A) UMAP colored by cluster identity: immature, mature, aged and interferon (IFN)-activated neutrophils. (B) Dot plot of the top four marker genes per cluster (Wilcoxon rank-sum test, ranked by log2 fold-change; adjusted p<0.05); dot color indicates average scaled expression, dot size indicates percentage of cells expressing each gene. (C) Over-representation analysis of Gene Ontology Biological Process terms of cluster marker genes; dot color indicates –log10(adjusted p), dot size indicates gene ratio. (D) Cluster composition (% of neutrophils per patient) by CMD status (CFR<2.5). The proportion of IFN-activated neutrophils was significantly different between the groups (p=0.048). (E-G) Percentage of IFN-activated neutrophils per patient versus CFR (E), systemic CD62L MFI measured by AQUIOS flow cytometry (F), and mean *SELL* expression (G). Dashed lines show the linear regression fit with 95% CI; rho denotes Spearman’s correlation. (H) Pseudotime trajectory analysis (Slingshot, rooted in the immature cluster), showing two lineages diverging after the mature stage towards the aged (1) and IFN-activated (2) fates; cells are colored by pseudotime. (I) Marker genes from (B), loess-smoothed and z-scored across pseudotime, for the aged and IFN lineage. (J) Median pseudotime along the IFN-activated lineage per patient vs. CFR; colored by CMD status and the dotted vertical line shows the CFR<2.5 threshold. Dashed line and rho as in (E-G).

The proportion of IFN-activated neutrophils was more than doubled in patients with a CMD diagnosis compared to those without a CMD diagnosis (median 17.1% vs. 6.3%, p=0.048, adj. p=0.0036) (**Figure 3D**). This was also true for the relation with CFR which proved to be inverse with CFR (rho=-0.74, p=0.013; β=-0.65, adj. p=0.087) (**Figure 3E**). The proportions of the other three neutrophil clusters were similar between patients with CMD and without CMD (all p>0.53).

Given that surface CD62L shedding is a well-established marker of neutrophil activation^39^, we examined whether the IFN-activated neutrophil state was associated with this activation phenotype by relating its proportion to neutrophil surface CD62L MFI in the same patients. We found that the proportion of IFN-activated neutrophils was inversely correlated with CD62L MFI (n=10, rho=-0.68, p=0.030; β=-0.67, adj. p=0.024) (**Figure 3F**) in ANOCA patients. In addition, *SELL* transcript expression (the gene encoding CD62L), was positively correlated with the proportion of IFN-activated neutrophils (n=11, rho = 0.74, p=0.013; β=0.77, adj. p=0.0083)) and negatively correlated with CFR (rho = −0.78, p=0.007; β= −0.71, adj. p=0.051) (**Figure 3G**). Together, these findings identify an IFN-activated neutrophil cluster that characterized both CMD and a lower CFR, and that was related to the shedding of CD62L from the cell surface. These findings were consistent in the women-only analysis, with similar trends for the associations between the IFN-activated cluster and CMD diagnosis, CFR, CD62L MFI and *SELL* expression (**Supplemental Figure S5**).

To assess whether the four clusters represent independent states or a transcriptional continuum, we performed pseudotime trajectory analysis, setting the immature neutrophil cluster as a starting point. This identified two trajectories originating from the immature cluster and passing through the mature cluster, one terminating in the IFN-activated cluster and the other in the aged cluster (**Figure 3H and 3I**). Within the lineage terminating in IFN-activated neutrophils, we identified that pseudotime position along the trajectory towards the IFN-activated fate (**trajectory 2, Figure 3H**) was further advanced in patients with a lower CFR (rho = −0.64, p=0.033) (**Figure 3J**). This supports that both the proportion of the IFN-activated neutrophils is higher and neutrophils overall are further along the IFN trajectory in patients with a lower CFR.

### 5. Immature neutrophils exhibit a distinct translational gene signature associated with low CFR suggestive of altered maturation

To investigate whether CFR-associated transcriptional changes were specific to particular neutrophil clusters, we performed cluster-specific pseudobulk differential expression analyses, modelling CFR as a continuous variable. We did not observe significant differentially expressed genes in the mature, the IFN-activated, or aged neutrophil clusters. In contrast, the immature cluster exhibited a distinct transcriptional signature associated with CFR. This comprised 140 differentially expressed genes (adjusted p <0.05, corrected for sequencing depth and cell number). This signature was dominated by genes encoding ribosomal proteins and translation machinery (e.g., *RPL9, RPL32, RPL6, RPS4X, RPL13, RPS17)* (**Figure 4A, Supplemental Table S8**). Gene set enrichment analysis showed that pathways indicating proliferation and biosynthesis (e.g., MYC targets, E2F targets, and oxidative phosphorylation) were lower in immature neutrophils from patients with a lower CFR measurement. Pathways reflecting cytokine-driven inflammatory signaling (e.g., IFN response and TNFα signaling) were higher in immature neutrophils from patients with a lower CFR measurement (**Figure 4B, Supplemental Table S9**). Consistent with this inflammatory phenotype, immature neutrophils from patients with a lower CFR also showed higher expression of *CXCR1* and *CXCR2*, which encode receptors for CXCL6, a chemokine that we recently found to be more abundant in the plasma of ANOCA patients with a lower CFR (**Supplemental Figure S6)** (Kregting, BHCM et al., unpublished data, 2026).

**Figure 4.**
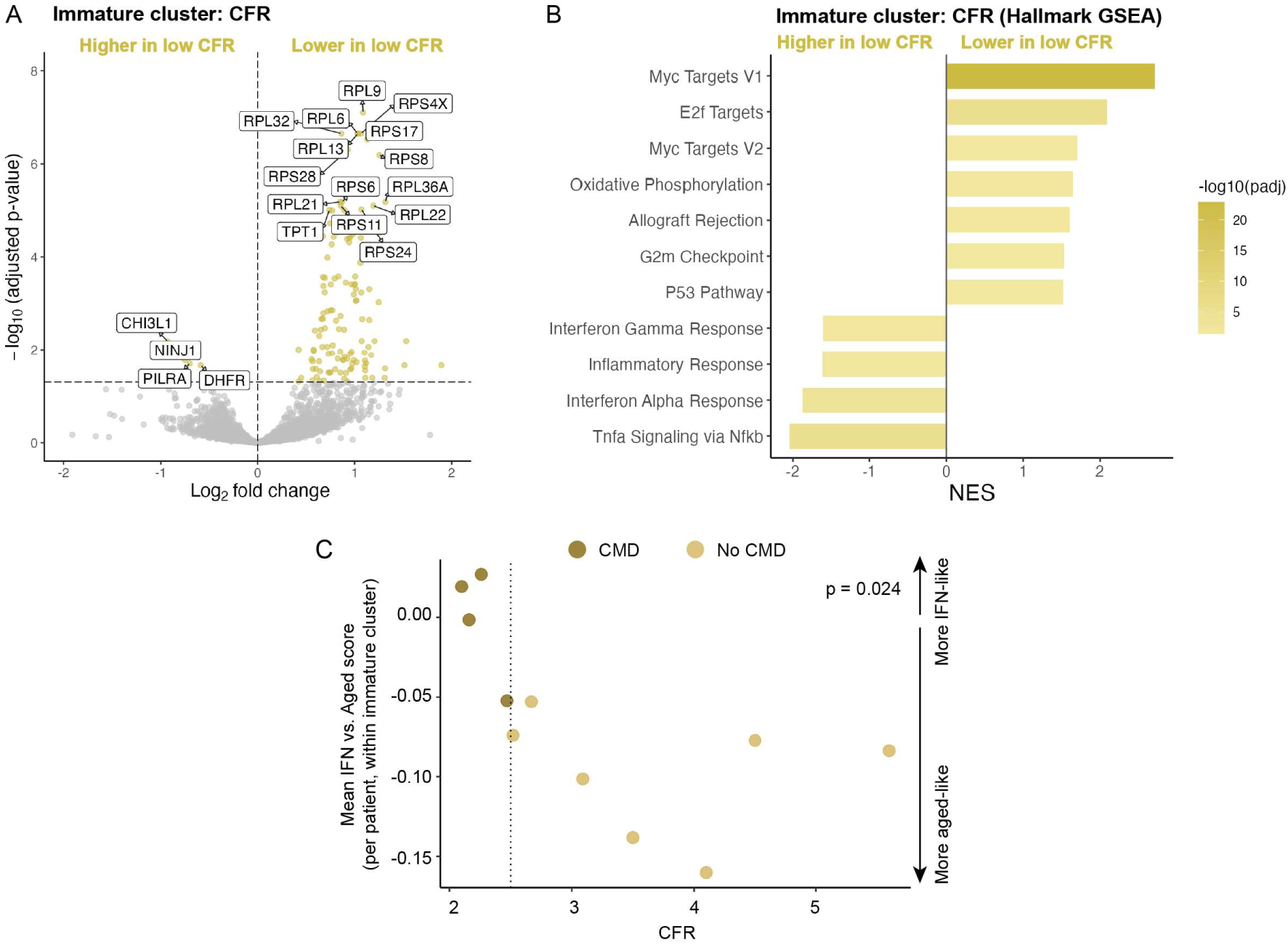
CFR-associated transcriptional signature in the immature neutrophil cluster. (A) Volcano plot of genes differentially expressed in the immature neutrophil cluster in association with coronary flow reserve (CFR, pseudobulk DESeq2, corrected for sequencing depth and cell number). Genes with adjusted p<0.05 are highlighted in yellow; selected top genes are labeled. (B) Hallmark gene set enrichment analysis (GSEA) of the CFR-associated signature shown in (A), ranked by DESeq2 Wald statistic. Bar color indicates –log10(adjusted p-value); NES, normalized enrichment score. (C) Mean interferon (IFN) minus aged-neutrophil module score per patient, calculated among cells within the immature neutrophil cluster, plotted against CFR. Points are colored by coronary microvascular dysfunction (CMD) diagnosis (CMD: CFR<2.5); the dotted line indicates the CFR<2.5 threshold. P-value from a linear mixed-effects model relating single-cell module score to CFR (p=0.04).

A parallel analysis using CMD diagnosis (CFR<2.5) as a binary variable showed similar results, with 83 of the 140 CFR-associated genes also significantly differentially expressed by CMD status (**Supplemental Table S9 and S10**). A sensitivity analysis restricted to women yielded a highly similar signature across both comparisons (**Supplemental Table S11-S14**). Thus, in addition to the higher percentage of IFN-activated neutrophils, lower CFR was associated with a distinct transcriptional phenotype in immature neutrophils marked by lower expression in ribosomal/translational and proliferative gene expression alongside increased inflammatory signaling. As IFN response signaling was enriched in immature neutrophils from patients with a lower CFR, we hypothesized that this reflects an early skewing towards the IFN-activated phenotype. We therefore asked whether immature neutrophils from patients with low CFR already resemble the IFN-activated cluster. Within the immature neutrophils, transcriptional profiles were leaning towards an IFN-activated (relative to an aged) phenotype in patients with a lower CFR (p=0.024, mixed-effects model accounting for multiple cells per patient) (**Figure 4C, Supplemental Table S15),** suggesting that immature neutrophils in patients with lower CFR are already transcriptionally primed towards an IFN-activated state.

### 6. Lower CD62L MFI on neutrophil surface is linked to altered endothelial transcriptional programs

In ANOCA patients, the IFN-activated neutrophil cluster we identified was associated with lower surface expression of CD62L, which is known to be relevant for neutrophil adhesion and migration^39^. We hypothesized that a lower CD62L MFI might be linked to endothelial cell states. To study this, we isolated coronary endothelial cells from catheterization material used during the diagnostic coronary function test in these patients (n=7, all women, **Supplemental Table S16**), as described before^24^. We performed bulk RNA sequencing and differential gene expression analysis using CD62L MFI as measured by the AQUIOS as a continuous variable.

We identified 31 genes that significantly increased (Log2FC <-0.25; e.g., *MT1X, MT2A, LDLR, MT1E, RSPO3, HMGCS1, CRIP1, FASN, ERAP2, ECI2*) and 161 genes that significantly decreased (Log2FC>0.25; e.g., *CCL14, TWIST2, SNED1, CLU, APOD, CXCL16, CORO2B, ACTA2, TSPAN15, AQP1*) with a lower CD62L MFI (adjusted p <0.05) (**Figure 5A, Supplemental Table S17**). Among these, *LDLR* encodes the LDL receptor, mediating cellular cholesterol uptake^40^, while *TWIST2* and *ACTA2* are established markers of endothelial-to-mesenchymal transition^41,42^. Gene set enrichment analysis (Hallmark) revealed significant positive enrichment of proliferative and metabolic pathways with a lower CD62L MFI (all adjusted p<0.05). Conversely, pathways reflecting IFN, complement and coagulation signaling, together with epithelial-to-mesenchymal transition and inflammatory/chemokine signaling, were negatively enriched with a lower CD62L MFI (all adjusted p<0.05) (**Figure 5B, Supplemental Table S18**). These findings suggest that a lower CD62L MFI is associated with a shift in endothelial gene expression, characterized by reduced expression of genes and pathways characteristic of an immune-competent, quiescent endothelial phenotype and increased expression of proliferative and metabolic genes.

**Figure 5.**
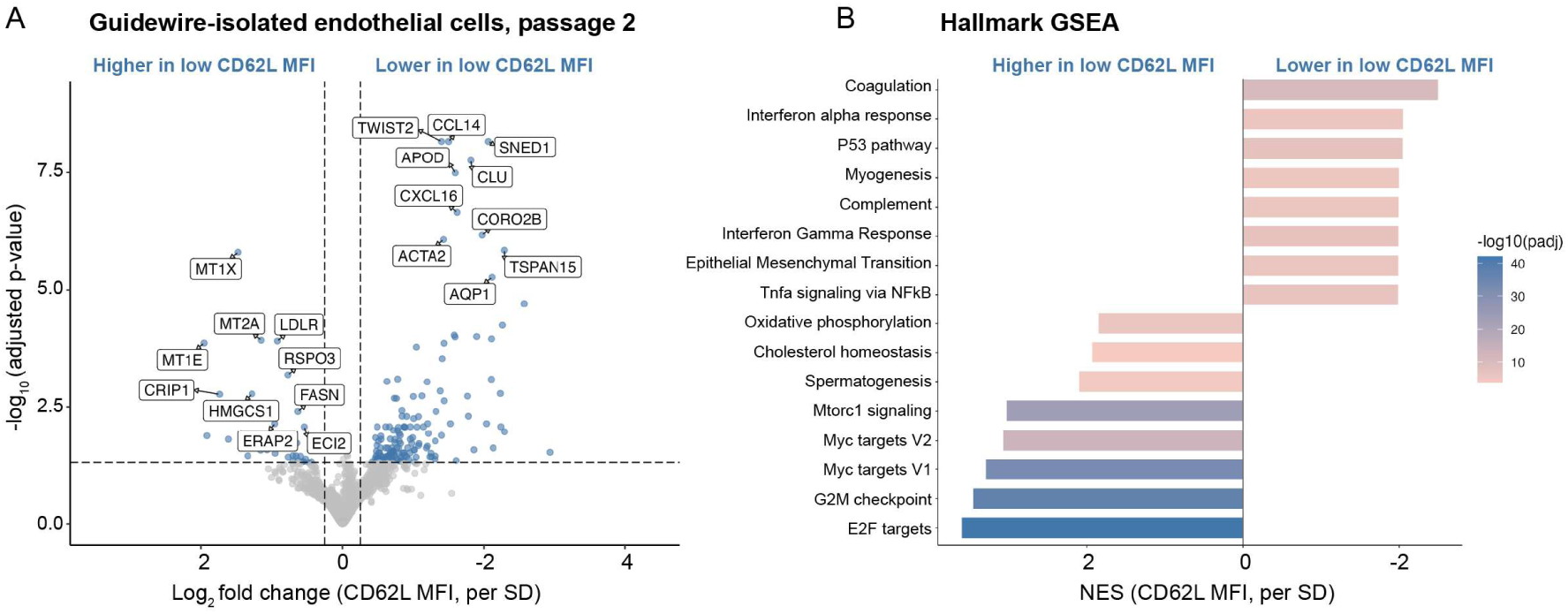
Guidewire-isolated endothelial cell transcriptional signature associated with neutrophil CD62L surface expression. Bulk RNA sequencing of guidewire-isolated endothelial cells (passage 2) from women with ANOCA, related to systemic neutrophil surface CD62L expression (AQUIOS flow cytometry). (A) Volcano plot of genes differentially expressed (DESeq2) per SD increase in CD62L median fluorescence intensity (MFI) (log2 fold change = ±0.25; age-adjusted p<0.05). Selected top genes are labeled. (B) Hallmark gene set enrichment analysis (GSEA) of the CD62L MFI-associated signature shown in (A), ranked by DESeq2 Wald statistic. Bar color indicates –log10(adjusted p); NES, normalized enrichment score (per SD CD62L MFI)

## Discussion

In this study, we characterized the circulating neutrophil phenotype in women with CMD and lower CFR using multiple complementary approaches. Women with coronary microvascular dysfunction (CMD) and lower coronary flow reserve (CFR) showed a distinct circulating neutrophil phenotype. Lower CFR was associated with higher neutrophil counts and inflammatory indices (NLR, SII), more activated CD62L^dim^ neutrophils, and a larger interferon (IFN)-activated neutrophil cluster identified by single-cell RNA sequencing. Immature neutrophils from these patients had reduced ribosomal and translational gene expression and were already skewed toward the IFN-activated profile, which may point to altered neutrophil maturation. In patient-derived endothelial cells, lower CD62L expression on circulating neutrophils was linked to a shift from an immune-supportive state toward greater proliferative and metabolic activity, suggesting these neutrophils may affect endothelial biology.

The pathophysiology of CMD remains incompletely understood. Previous studies on microvascular diseases have primarily focused on circulating neutrophil counts and NETosis^18,19,35,43^, providing limited insight into neutrophil activation and transcriptional heterogeneity. By combining complete blood counts, flow cytometric activation phenotyping and single-cell transcriptomics, our study provides a multidimensional characterization of circulating neutrophils in relation to CMD. Importantly, these analyses were performed in a well-phenotyped clinical cohort with invasively measured CFR, enabling assessment of the relationship between neutrophil phenotype and an objective, quantitative measure of coronary microvascular function.

The female predominance of CMD raises the question whether sex-specific immune biology contributes to the neutrophil phenotype observed in our study. Indeed, neutrophils from healthy young women show stronger type I IFN signatures than those from men, despite similar circulating IFN activity and IFN receptor expression. This difference appears to reflect enhanced intracellular signaling, including greater nuclear localization of IFN-responsive transcription factors, rather than greater IFN exposure. The female-biased response was linked to estrogen and was absent before puberty, suggesting that sex hormones may shape neutrophil responsiveness^44^. Complementing these observations, a recent single-cell atlas of human neutrophils identified a higher proportion of neutrophils expressing combined type I and II IFN-associated programs in healthy female donors^33^. Together, these observations suggest that the neutrophil phenotype observed in our CMD cohort reflects, in part, an amplification of pre-existing sex differences in neutrophil IFN responsiveness. Related evidence of sex-specific IFN-associated pathways in cardiovascular disease, including heart failure with preserved ejection fraction^45^ and other female-predominant conditions^46,47^, further supports the potential relevance of sex-dependent IFN signaling in shaping immune phenotypes and contributing to disease susceptibility. This raises the possibility that these differences precede clinical disease, although longitudinal studies are needed to establish their contribution to CMD.

Importantly, our findings suggest that this IFN-biased phenotype may emerge already early in neutrophil maturation. Our pseudotime analysis indicated that immature neutrophils from patients with lower CFR were already transcriptionally skewed towards an IFN-activated state. Consistent with this, the association between CD62L^dim^ neutrophils and CFR observed at baseline was no longer evident following *ex vivo* fNLF stimulation. Although stimulation may alter CD62L surface expression and obscure baseline differences, this finding suggests that lower CFR is associated with a pre-existing neutrophil state that is not simply explained by an enhanced acute response to stimulation.

Endothelial dysfunction contributes to impaired coronary microvascular regulation in CMD^3^. The proportion of IFN-activated neutrophils identified in our study was associated with a reduced surface CD62L, pointing to altered neutrophil-endothelial interactions. CD62L mediates initial neutrophil rolling, and its shedding accompanies activation and the transition to firm adhesion^39^. Despite lower surface CD62L, *SELL* transcript expression was higher with increasing proportions of IFN-activated neutrophils, potentially reflecting ongoing transcription alongside protein shedding. However, our data cannot establish whether shedding is ongoing or whether these neutrophils interact directly with the endothelium. In our endothelial cell data, we found that lower neutrophil CD62L MFI was associated with increased endothelial proliferative and metabolic activity, consistent with a shift away from a quiescent state. This may be consistent with evidence that type I IFN signaling can promote endothelial activation and leukocyte recruitment, reduce nitric oxide bioavailability, impair endothelial repair and promote NET formation and thrombosis^48–50^. IFN-activated neutrophils may therefore influence endothelial function through altered adhesion or inflammatory signaling, potentially contributing to microvascular dysfunction. However, the direction of this relationship remains unclear: the neutrophil phenotype may contribute to endothelial dysfunction, arise in response to pre-existing abnormalities, or reflect a shared underlying process. These findings raise the possibility that IFN-associated neutrophil activation may be a therapeutic target in CMD, although further functional studies are needed to establish its causal role and therapeutic relevance.

Future work should build on our associative findings in several directions. Longitudinal studies are needed to determine whether this neutrophil phenotype precedes the development of CMD or arises as a consequence of reduced CFR. Some of our data suggests that, at least in part, this phenotype might be encoded already in immature neutrophil stages. Functional studies are needed to establish whether CD62L shedding and the IFN-activated neutrophil phenotype causally affect neutrophil-endothelial adhesion and coronary microvascular function. In addition, the higher *CXCR1* and *CXCR2* expression in immature neutrophils from patients with a lower CFR, together with our parallel observation of elevated plasma CXCL6 in patients with a lower CFR, suggests a candidate signaling axis that merits further mechanistic investigation (Kregting, BHCM et al., unpublished data, 2026). It also remains unclear whether this circulating phenotype preferentially affects the coronary microcirculation or instead, reflects a more generalized marker of systemic microvascular vulnerability. Examining whether specific features of the coronary endothelium, or its hemodynamic environment confer susceptibility, would help clarify this. Finally, direct measurements of circulating IFN levels and sex-hormones would allow the proposed estrogen-driven mechanism of neutrophil IFN predisposition to be tested directly, rather than inferred from prior literature in healthy cohorts.

This study has several limitations. First, the cross-sectional design precludes conclusions about causality, as discussed above. Second, this was a single-center study, which may limit generalizability. Third, the number of men included for single-cell RNA sequencing and the supplementary subgroup analyses was small, limiting statistical power for these analyses.

In conclusion, this study provides the first detailed phenotypic characterization of circulating neutrophils in patients with CMD. We identified a circulating neutrophil phenotype marked by an increase in IFN-activated neutrophils, an IFN-enriched immature neutrophil phenotype and CD62L shedding, with potential relevance to coronary endothelial function. These findings provide pathophysiological insight into a condition that disproportionately affects women, positioning neutrophil activation as a candidate contributor to CMD and underscoring the importance of sex-specific research in cardiovascular disease.

## Abbreviations

ANOCA: angina with non-obstructive coronary arteries
CFR: coronary flow reserve
CMD: coronary microvascular dysfunction
fNLF: N-formyl-norleucyl-leucyl-phenalalanine
IFN: interferon
MFI: median fluorescent intensity
NETs: neutrophil extracellular traps
NT-proBNP: N-terminal pro-B-type natriuretic peptide

## Data Availability

Data supporting the findings of this study are available from the corresponding author upon reasonable request.

## References

1. Samuels BA, Shah SM, Widmer RJ, Kobayashi Y, Miner SES, Taqueti VR, Jeremias A, Albadri A, Blair JA, Kearney KE, et al. Comprehensive Management of ANOCA, Part 1-Definition, Patient Population, and Diagnosis: JACC State-of-the-Art Review. J Am Coll Cardiol. 2023;82:1245–1263. doi: 10.1016/j.jacc.2023.06.043

2. Kunadian V, Chieffo A, Camici PG, Berry C, Escaned J, Maas A, Prescott E, Karam N, Appelman Y, Fraccaro C, et al. An EAPCI Expert Consensus Document on Ischaemia with Non-Obstructive Coronary Arteries in Collaboration with European Society of Cardiology Working Group on Coronary Pathophysiology & Microcirculation Endorsed by Coronary Vasomotor Disorders International Study Group. EuroIntervention. 2021;16:1049–1069. doi: 10.4244/EIJY20M07_01

3. Crea F, Montone RA, Rinaldi R. Pathophysiology of Coronary Microvascular Dysfunction. Circ J. 2022;86:1319–1328. doi: 10.1253/circj.CJ-21-0848

4. Taqueti VR, Solomon SD, Shah AM, Desai AS, Groarke JD, Osborne MT, Hainer J, Bibbo CF, Dorbala S, Blankstein R, et al. Coronary microvascular dysfunction and future risk of heart failure with preserved ejection fraction. Eur Heart J. 2018;39:840–849. doi: 10.1093/eurheartj/ehx721

5. Crooijmans C, Adriaansen FW, Meeder JG, Beijk MAM, Yosofi B, Appelman Y, Mechroubi A, de Vos A, Paradies V, Meuwissen M, et al. Patients with angina and non-obstructive coronary artery disease undergoing comprehensive invasive coronary function testing: long-term clinical outcomes. Eur Heart J. 2026. doi: 10.1093/eurheartj/ehag649

6. Del Buono MG, Montone RA, Camilli M, Carbone S, Narula J, Lavie CJ, Niccoli G, Crea F. Coronary Microvascular Dysfunction Across the Spectrum of Cardiovascular Diseases: JACC State-of-the-Art Review. J Am Coll Cardiol. 2021;78:1352–1371. doi: 10.1016/j.jacc.2021.07.042

7. Dal Canto E, Diez Benavente E, van de Hoef T, Bouatia-Naji N, Reue K, Civelek M, den Ruijter H. Redesigning cardiovascular medicine around sex differences. Nat Med. 2026. doi: 10.1038/s41591-026-04659-1

8. Smilowitz NR, Schlamp F, Hausvater A, Joa A, Serrano-Gomez C, Farid A, Hochman JS, Barrett TJ, Reynolds HR, Berger JS. Coronary Microvascular Dysfunction Is Associated With a Proinflammatory Circulating Transcriptome in Patients With Nonobstructive Coronary Arteries. Arterioscler Thromb Vasc Biol. 2024;44:997–999. doi: 10.1161/ATVBAHA.123.320471

9. Guo Z, Yang Z, Song Z, Li Z, Xiao Y, Zhang Y, Wen T, Pan G, Xu H, Sheng X, et al. Inflammation and coronary microvascular disease: relationship, mechanism and treatment. Front Cardiovasc Med. 2024;11:1280734. doi: 10.3389/fcvm.2024.1280734

10. Tona F, Serra R, Di Ascenzo L, Osto E, Scarda A, Fabris R, Montisci R, Famoso G, Tellatin S, Foletto M, et al. Systemic inflammation is related to coronary microvascular dysfunction in obese patients without obstructive coronary disease. Nutr Metab Cardiovasc Dis. 2014;24:447–453. doi: 10.1016/j.numecd.2013.09.021

11. Recio-Mayoral A, Mason JC, Kaski JC, Rubens MB, Harari OA, Camici PG. Chronic inflammation and coronary microvascular dysfunction in patients without risk factors for coronary artery disease. Eur Heart J. 2009;30:1837–1843. doi: 10.1093/eurheartj/ehp205

12. Weber B, Weisenfeld D, Massarotti E, Seyok T, Cremone G, Lam E, Golnik C, Brownmiller S, Liu F, Huang S, et al. Interplay Between Systemic Inflammation, Myocardial Injury, and Coronary Microvascular Dysfunction in Rheumatoid Arthritis: Results From the LiiRA Study. J Am Heart Assoc. 2024;13:e030387. doi: 10.1161/JAHA.123.030387

13. Weber B, Weisenfeld D, Seyok T, Huang S, Massarotti E, Barrett L, Bibbo C, Solomon DH, Plutzky J, Bolster M, et al. Relationship Between Risk of Atherosclerotic Cardiovascular Disease, Inflammation, and Coronary Microvascular Dysfunction in Rheumatoid Arthritis. J Am Heart Assoc. 2022;11:e025467. doi: 10.1161/JAHA.121.025467

14. Weber BN, Stevens E, Barrett L, Bay C, Sinnette C, Brown JM, Divakaran S, Bibbo C, Hainer J, Dorbala S, et al. Coronary Microvascular Dysfunction in Systemic Lupus Erythematosus. J Am Heart Assoc. 2021;10:e018555. doi: 10.1161/JAHA.120.018555

15. Ambler WG, Kaplan MJ. Vascular damage in systemic lupus erythematosus. Nat Rev Nephrol. 2024;20:251–265. doi: 10.1038/s41581-023-00797-8

16. Wirestam L, Arve S, Linge P, Bengtsson AA. Neutrophils-Important Communicators in Systemic Lupus Erythematosus and Antiphospholipid Syndrome. Front Immunol. 2019;10:2734. doi: 10.3389/fimmu.2019.02734

17. Jariwala MP, Laxer RM. NETosis in Rheumatic Diseases. Curr Rheumatol Rep. 2021;23:9. doi: 10.1007/s11926-020-00977-6

18. Levit R, Wang L, Gaignard S, Sayegh M, Vatsa N, Rashid FA, McDowell A, Mehta PK. NEUTROPHILS AND NETOSIS IN WOMEN WITH CORONARY MICROVASCULAR DYSFUNCTION: A HYPOTHESIS GENERATING STUDY. JACC. 2022;79:992–992. doi: doi:10.1016/S0735-1097(22)01983-0

19. Shirakawa K, Sano M. Neutrophils and Neutrophil Extracellular Traps in Cardiovascular Disease: An Overview and Potential Therapeutic Approaches. Biomedicines. 2022;10. doi: 10.3390/biomedicines10081850

20. Meegan JE, Yang X, Coleman DC, Jannaway M, Yuan SY. Neutrophil-mediated vascular barrier injury: Role of neutrophil extracellular traps. Microcirculation. 2017;24. doi: 10.1111/micc.12352

21. Meteva D, Vinci R, Seppelt C, Abdelwahed YS, Pedicino D, Nelles G, Skurk C, Haghikia A, Rauch-Krohnert U, Gerhardt T, et al. Toll-like receptor 2, hyaluronan, and neutrophils play a key role in plaque erosion: the OPTICO-ACS study. Eur Heart J. 2023;44:3892–3907. doi: 10.1093/eurheartj/ehad379

22. Meteva D, Pedicino D, Abdelwahed YS, Krankel N, Anwari E, Leistner DM, den Ruijter H, Gerhardt T, Joner M, Libby P, et al. Acute coronary syndrome with plaque erosion: clinical characteristics, mechanisms, and unmet needs. Eur Heart J. 2026;47:1403–1414. doi: 10.1093/eurheartj/ehaf1004

23. Crooijmans C, Jansen TPJ, Konst RE, Woudstra J, Appelman Y, den Ruijter HM, Onland-Moret NC, Meeder JG, de Vos AMJ, Paradies V, et al. Design and rationale of the NetherLands registry of invasive Coronary vasomotor Function Testing (NL-CFT). Int J Cardiol. 2023;379:1–8. doi: 10.1016/j.ijcard.2023.02.043

24. de Jong EAM, Kapteijn DMC, Daniels MA, Nijkamp T, Zalewski PD, Beltrame JF, Damman P, Civelek M, Diez Benavente E, van de Hoef TP, et al. Feasibility of endothelial cell isolation from routine coronary function testing in ANOCA patients. Int J Cardiol. 2026;462:134711. doi: 10.1016/j.ijcard.2026.134711

25. Ong P, Camici PG, Beltrame JF, Crea F, Shimokawa H, Sechtem U, Kaski JC, Bairey Merz CN, Coronary Vasomotion Disorders International Study G. International standardization of diagnostic criteria for microvascular angina. Int J Cardiol. 2018;250:16–20. doi: 10.1016/j.ijcard.2017.08.068

26. De Bruyne B, Pijls NHJ, Gallinoro E, Candreva A, Fournier S, Keulards DCJ, Sonck J, Van’t Veer M, Barbato E, Bartunek J, et al. Microvascular Resistance Reserve for Assessment of Coronary Microvascular Function: JACC Technology Corner. J Am Coll Cardiol. 2021;78:1541–1549. doi: 10.1016/j.jacc.2021.08.017

27. Gallinoro E, Bertolone DT, Mizukami T, Paolisso P, Bermpeis K, Munhoz D, Sakai K, Seki R, Ohashi H, Esposito G, et al. Continuous vs Bolus Thermodilution to Assess Microvascular Resistance Reserve. JACC Cardiovasc Interv. 2023;16:2767–2777. doi: 10.1016/j.jcin.2023.09.027

28. ten Berg MJ, Huisman A, van den Bemt PM, Schobben AF, Egberts AC, van Solinge WW. Linking laboratory and medication data: new opportunities for pharmacoepidemiological research. Clin Chem Lab Med. 2007;45:13–19. doi: 10.1515/CCLM.2007.009

29. Spijkerman R, Hesselink L, Hellebrekers P, Vrisekoop N, Hietbrink F, Leenen LPH, Koenderman L. Automated flow cytometry enables high performance point-of-care analysis of leukocyte phenotypes. J Immunol Methods. 2019;474:112646. doi: 10.1016/j.jim.2019.112646

30. de Fraiture EJ, Bongers SH, Jukema BN, Koenderman L, Vrisekoop N, van Wessem KJP, Leenen LPH, Hietbrink F. Visualization of the inflammatory response to injury by neutrophil phenotype categories: Neutrophil phenotypes after trauma. Eur J Trauma Emerg Surg. 2023;49:1023–1034. doi: 10.1007/s00068-022-02134-3

31. Spijkerman R, Jorritsma NKN, Bongers SH, Bindels BJJ, Jukema BN, Hesselink L, Hietbrink F, Leenen LPH, van Goor HMR, Vrisekoop N, et al. An increase in CD62L(dim) neutrophils precedes the development of pulmonary embolisms in COVID-19 patients. Scand J Immunol. 2021;93:e13023. doi: 10.1111/sji.13023

32. Singleron Biotechnologies. GEXSCOPE Single Cell RNA Library Kit for NEO: Handbook (NEO Chip SD/HD - 4 RXNs/16 RXNs). In: Cologne, Germany: Singleron Biotechnologies GmbH; 2025.

33. Zheng H, Maestri E, Moore AR, Ganesan A, Enriquez J, Duran V, Einav S, van der Poll T, Bouma HR, Atreya MR, et al. A single-cell atlas of human neutrophils reveals neutrophil heterogeneity across health and disease. Nat Immunol. 2026;27:2322–2334. doi: 10.1038/s41590-026-02671-8

34. Street K, Risso D, Fletcher RB, Das D, Ngai J, Yosef N, Purdom E, Dudoit S. Slingshot: cell lineage and pseudotime inference for single-cell transcriptomics. BMC Genomics. 2018;19:477. doi: 10.1186/s12864-018-4772-0

35. Shi X, Sun J, Wang H, Zhao Q, Yan C, Cao Y. Combined diagnostic value of the systemic immune-inflammation index and LDL for coronary microvascular disease: a retrospective cohort study. Ther Adv Cardiovasc Dis. 2026;20:17539447261469508. doi: 10.1177/17539447261469508

36. Merdler I, Chitturi KR, Chaturvedi A, Li J, Cellamare M, Ozturk ST, Sawant V, Ben-Dor I, Waksman R, Case BC, et al. Coronary microvascular dysfunction and inflammation: Insights from the Coronary Microvascular Disease Registry. Cardiovasc Revasc Med. 2024;69:30–34. doi: 10.1016/j.carrev.2024.05.020

37. Karakayali M, Altunova M, Yakisan T, Aslan S, Omar T, Artac I, Ilis D, Arslan A, Cagin Z, Karabag Y, et al. The Relationship between the Systemic Immune-Inflammation Index and Ischemia with Non-Obstructive Coronary Arteries in Patients Undergoing Coronary Angiography. Arq Bras Cardiol. 2024;121:e20230540. doi: 10.36660/abc.20230540

38. Kalhor P, Mahmoudi Hamidabad N, Hellou E, Nogami K, Manzato M, Lerman LO, Lerman A. Association of monocyte-to-lymphocyte ratio with coronary endothelial dysfunction and cardiovascular events in patients with angina and nonobstructive coronary artery disease. Atherosclerosis. 2025;410:120539. doi: 10.1016/j.atherosclerosis.2025.120539

39. Connelly AN, Huijbregts RPH, Pal HC, Kuznetsova V, Davis MD, Ong KL, Fay CX, Greene ME, Overton ET, Hel Z. Optimization of methods for the accurate characterization of whole blood neutrophils. Sci Rep. 2022;12:3667. doi: 10.1038/s41598-022-07455-2

40. Go GW, Mani A. Low-density lipoprotein receptor (LDLR) family orchestrates cholesterol homeostasis. Yale J Biol Med. 2012;85:19–28.

41. Yoshimatsu Y, Watabe T. Emerging roles of inflammation-mediated endothelial-mesenchymal transition in health and disease. Inflamm Regen. 2022;42:9. doi: 10.1186/s41232-021-00186-3

42. Sapoznik KA, Gast TJ, Carmichael-Martins A, Walker BR, Warner RL, Burns SA. Retinal Arteriolar Wall Remodeling in Diabetes Captured With AOSLO. Transl Vis Sci Technol. 2023;12:16. doi: 10.1167/tvst.12.11.16

43. Chakrala T, Prakash R, Valdes C, Pepine CJ, Keeley EC. Circulating Biomarkers in Coronary Microvascular Dysfunction. J Am Heart Assoc. 2023;12:e029341. doi: 10.1161/JAHA.122.029341

44. Gupta S, Nakabo S, Blanco LP, O’Neil LJ, Wigerblad G, Goel RR, Mistry P, Jiang K, Carmona-Rivera C, Chan DW, et al. Sex differences in neutrophil biology modulate response to type I interferons and immunometabolism. Proc Natl Acad Sci U S A. 2020;117:16481–16491. doi: 10.1073/pnas.2003603117

45. van Ommen AM, Diez Benavente E, Onland-Moret NC, Valstar GB, Cramer MJ, Rutten FH, Teske AJ, Menken R, Hofstra L, Tulevski, II, et al. Plasma Proteomic Patterns Show Sex Differences in Early Concentric Left Ventricular Remodeling. Circ Heart Fail. 2023;16:e010255. doi: 10.1161/CIRCHEARTFAILURE.122.010255

46. Gupta S, Kaplan MJ. Bite of the wolf: innate immune responses propagate autoimmunity in lupus. J Clin Invest. 2021;131. doi: 10.1172/JCI144918

47. Nocturne G, Mariette X. Interferon signature in systemic autoimmune diseases: what does it mean? RMD Open. 2022;8. doi: 10.1136/rmdopen-2022-002687

48. Nishide M, Nishimura K, Matsushita H, Kawada S, Shimagami H, Metsugi S, Kato Y, Kawasaki T, Tsujimoto K, Edahiro R, et al. Neutrophil single-cell analysis identifies a type II interferon-related subset for predicting relapse of autoimmune small vessel vasculitis. Nat Commun. 2025;16:3581. doi: 10.1038/s41467-025-58550-7

49. Thacker SG, Zhao W, Smith CK, Luo W, Wang H, Vivekanandan-Giri A, Rabquer BJ, Koch AE, Pennathur S, Davidson A, et al. Type I interferons modulate vascular function, repair, thrombosis, and plaque progression in murine models of lupus and atherosclerosis. Arthritis Rheum. 2012;64:2975–2985. doi: 10.1002/art.34504

50. Somers EC, Zhao W, Lewis EE, Wang L, Wing JJ, Sundaram B, Kazerooni EA, McCune WJ, Kaplan MJ. Type I interferons are associated with subclinical markers of cardiovascular disease in a cohort of systemic lupus erythematosus patients. PLoS One. 2012;7:e37000. doi: 10.1371/journal.pone.0037000

